# Coronavirus Disease 2019 (COVID-19) Candidate Chest CT Features: A Systematic Review of Extracted Imaging Features from 7571 Individuals

**DOI:** 10.1101/2020.11.03.20225326

**Authors:** Javad Zahiri, Mohammad Hossein Afsharinia, Zhaleh Hekmati, Mohsen Khodarahmi, Shahrzad Hekmati, Ramin Pourghorban

**Affiliations:** Bioinformatics and Computational Omics Lab (BioCOOL), Department of Biophysics, Faculty of Biological Sciences, Tarbiat Modares University, Tehran, Iran; Dr Khodarahmi Medical Imaging Center, Karaj, Iran; Department of Radiology, Madaran Hospital, Tehran, Iran; Department of Radiology, Sina Hospital, EBIR Tehran University of Medical Sciences, Tehran, Iran

**Keywords:** CT, chest computed tomography, imaging features, COVID-19, data mining, SARS, MERS

## Abstract

Since the outbreak of Coronavirus Disease 2019 (COVID-19) causing novel coronavirus (2019-nCoV)-infected pneumonia (NCIP), over 45 million affected cases have been reported worldwide. Many patients with COVID-19 have involvement of their respiratory system. According to studies in the radiology literature, chest computed tomography (CT) is recommended in suspected cases for initial detection, evaluating the disease progression and monitoring the response to therapy. The aim of this article is to review the most frequently reported imaging features in COVID-19 patients in order to provide a reliable insight into expected CT imaging manifestations in patients with positive reverse-transcription polymerase chain reaction (RT-PCR) test results, and also for the initial detection of patients with suspicious clinical presentation whose RT-PCR test results are false negative. A total of 60 out of 173 initial COVID-19 studies, comprising 7571 individuals, were identified by searching PubMed database for articles published between the months of January and June 2020. The data of these studies were related to patients from China, Japan, Italy, USA, Iran and Singapore. Among 40 reported features, presence of ground glass opacities (GGO), consolidation, bilateral lung involvement and peripheral distribution are the most frequently observed ones, reported in 100%, 91.7%, 85%, and 83.3% of articles, respectively. In a similar way, we extracted CT imaging studies of similar pulmonary syndromes outbreaks caused by other strains of coronavirus family: Middle East Respiratory Syndrome (MERS) and Severe Acute Respiratory Syndrome (SARS). For MERS and SARS, 2 out of 21 and 5 out of 153 initially retrieved studies had CT findings, respectively. Herein, we have indicated the most common coronavirus family related and COVID-19 specific features. Presence of GGO, consolidation, bilateral lung involvement and peripheral distribution were the features reported in at least 83% of COVID-19 articles, while air bronchogram, multi-lobe involvement and linear opacity were the three potential COVID-19 specific CT imaging findings. This is necessary to recognize the most promising imaging features for diagnosis and follow-up of patients with COVID-19. Furthermore, we identified co-existed CT imaging features.

## Introduction

In December 2019, a cluster of patients with pneumonia symptoms were reported in Wuhan city, Hubei Province, China. The etiology of illness remained unknown until a novel strain of coronavirus was determined to be responsible for outbreak. It was later named by World Health Organization (WHO) as the 2019-novel coronavirus (2019-nCoV)(1). Coronavirus disease 2019 (COVID-19) soon spread across continents and finally on March 11, 2020 WHO characterized it as a pandemic(2). Since ACE2, the entry receptor for the causative 2019-nCoV, is expressed in multiple tissues, it can result in several manifestations including thrombotic complications, myocardial dysfunction and arrhythmia, acute coronary syndromes, acute kidney injury, gastrointestinal symptoms, hepatocellular injury, hyperglycemia and ketosis, neurologic illnesses, ocular symptoms, and dermatologic complications(3). Despite all this, COVID-19 is most well-known for causing substantial respiratory pathology.

Due to lung damage in COVID-19, patients have typical CT imaging features that can be helpful in early screening of highly suspected cases and in evaluation of the severity and extent of disease(4). So it is important for all radiologists to be aware of the imaging spectrum of the disease and contribute to effective surveillance and response measures(5).

In Addition to having a pivotal role in evaluating the disease progression and monitoring the response to therapy, chest CT has a low rate of missed diagnosis of COVID-19 and may be useful as a standard method for the rapid diagnosis of COVID-19 to optimize the management of patients(6). However, viral nucleic acid detection with real-time polymerase chain reaction (RT-PCR) test remains the gold standard. Although chest CT may be negative for patients with COVID-19 at initial state of disease(7), many studies indicated that CT imaging has more sensitivity than RT-PCR(8). Therefore, CT is also recommended for the initial detection of patients with suspicious clinical presentation whose RT-PCR test results are false negative. Furthermore, Chest CT is recommended for follow-up in cases who are recovering from COVID-19 to evaluate permanent or long-term lung damage including fibrosis.

Features in COVID-19 CT images are variable and have significant overlap with those of Middle Eastern Respiratory Syndrome (MERS) and Severe Acute Respiratory Syndrome (SARS)(9). Therefore, we analyzed the reported CT findings from 67 different studies to identify the most frequent features within COVID-19 studies and also COVID-19 specific features. Given that the other types of viral pneumonia have similar radiological appearances with COVID-19, their differentiation is useful in facilitating the screening process in clinical practice(10).

In this study, we aim to identify the most important CT imaging features of patients with COVID-19, and then check whether there are some COVID-19 specific CT findings. Also, we examined the co-occurrence of features in the analyzed studies.

## Methods

### Data gathering

Figure 1 schematically shows how the data were gathered from the literature. Firstly, we performed a search in titles and abstracts of articles stored in Pubmed database using the keywords “COVID-19”,”SARS-CoV-2”,”SARS2” and “severe acute respiratory syndrome coronavirus 2” for COVID-19; “MERS”, “Middle East Respiratory Syndrome”, “MERS-coronavirus” or “MERS-CoV” for MERS; and “severe acute respiratory syndrome”, “SARS”, “SARS-CoV-1” and “SARSr-CoV” for SARS, each together with “chest CT” or “Computed Tomography” in order to obtain articles related to both coronavirus family and CT imaging. This search was resulted in 173, 153 and 21 articles for COVID-19, SARS and MERS, respectively.

**Figure 1.**
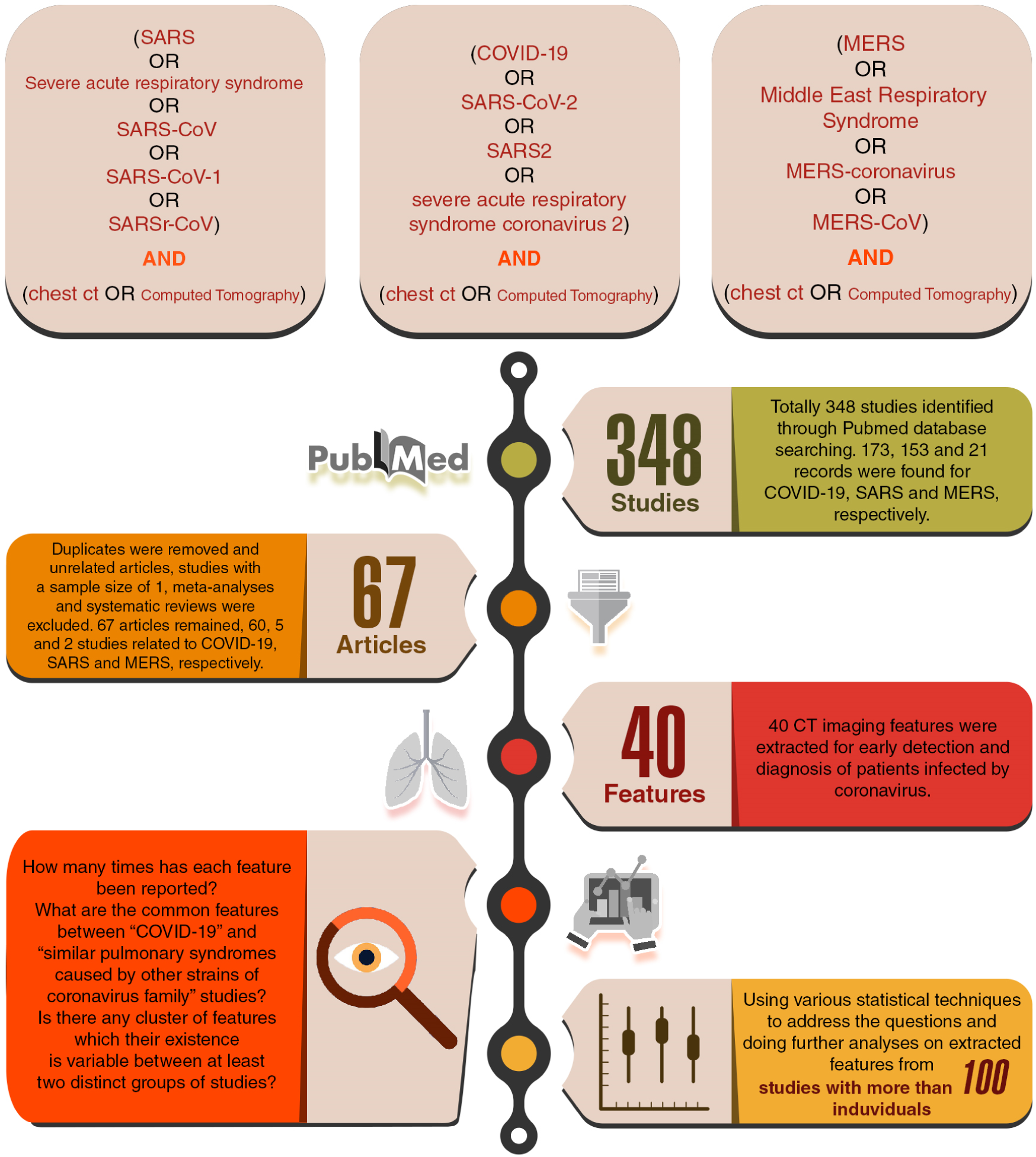
Schematic flowchart of this study

Then, duplications and unrelated articles (studies with a sample size of 1, studies with no reported CT findings, meta-analyses and systematic reviews) were removed. We finally achieved 60, 5 and 2 studies (comprising 7835 total individuals), reporting CT imaging features for COVID-19, SARS and MERS, respectively.

Figure 2 depicts the sample size distribution of all 67 studies for the three viral diseases. Among 60 COVID-19 articles, there were 19 studies which had more than 100 samples.

**Figure 2.**
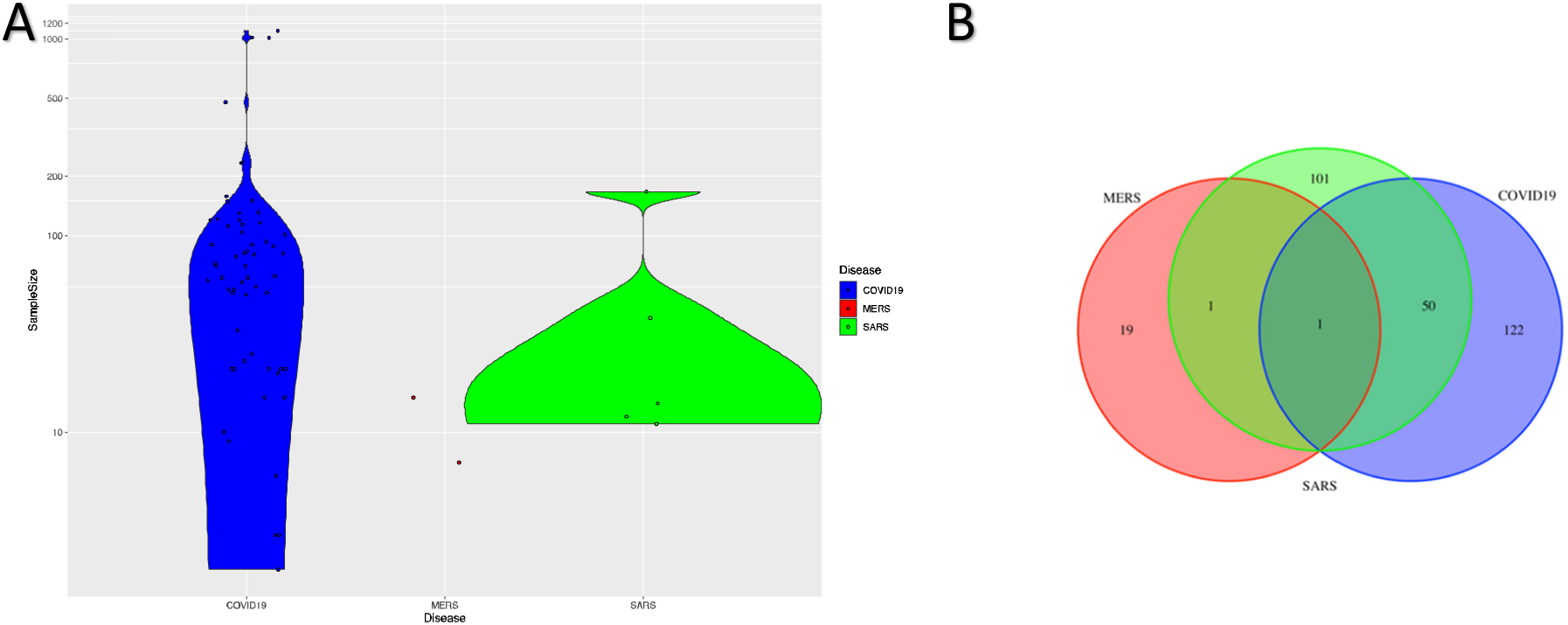
A. Violin plot comparing distribution of sample sizes of studies related to each coronavirus related disease. B. Venn diagram showing initial studies shared by COVID-19, SARS and MERS.

### CT features

We extracted 40 unique CT imaging features from the articles. Table 1 shows these features. Generally speaking, as Table 1 shows, these 40 CT features can be considered as three feature groups: lung involvement, main features and other features. Lung involvement includes 15 features which were categorized into three subgroups based on location, distribution and lesion. Main features include 6 features consisting of presence of GGO, morphology of GGO, consolidation, linear opacity, reticular opacity and septal thickening. Finally, the rest 19 CT features were considered as the “other features”.

**Table 1:** Categorized radiological findings on chest CT images of 7571 individuals of 60 COVID-19 studies.

|  |  |  |
| --- | --- | --- |
| Lung involvement | Location | Unilateral |
|  |  | Bilateral |
|  | Distribution | Upper |
|  |  | Middle |
|  |  | Lower |
|  |  | Central |
|  |  | Subpleural distribution |
|  |  | Peripheral |
|  |  | Central + peripheral |
|  |  | Anterior* |
|  |  | Posterior* |
|  | Lesion | Single Lesion |
|  |  | Multiple Lesions |
|  |  | Multilobe involvement |
|  |  | Diffuse |
| Main Features | Ground Glass Opacities (GGO) | Presence |
|  |  | Morphology* (rounded or not rounded) |
|  | Consolidation |  |
|  | Linear Opacity |  |
|  | Reticular Opacity |  |
|  | Septal Thickening** |  |
| Other Features | Air Bronchogram |  |
|  | Bronchial wall thickening |  |
|  | Bronchiectasis |  |
|  | Crazy-paving pattern |  |
|  | Halo Sign |  |
|  | Honeycomb pattern |  |
|  | Lymphadenopathy |  |
|  | Mucoid Impaction |  |
|  | Multifocal organizing pneumonia |  |
|  | Nodule |  |
|  | Pericardial Effusion |  |
|  | Pleural Effusion |  |
|  | Pleural Thickening |  |
|  | Pneumothorax |  |
|  | Pulmonary Cavities |  |
|  | Reverse Halo Sign |  |
|  | Spider web sign |  |
|  | Tree in bud sign |  |
|  | Vascular Thickening |  |

\* Many studies did not mention these detailed features (anterior, posterior and morphology of GGO).
\*\* For further analysis in the rest of this study, *septal thickening* was considered as the same as *reticular opacity*.

## Results and Discussion

According to the analyzed studies, some of the CT findings were reported more frequently in studies. Figure 3 shows the frequency of each CT feature in all COVID-19 studies. In the following, the 4 most frequent features are discussed in more details.

**Figure 3.**
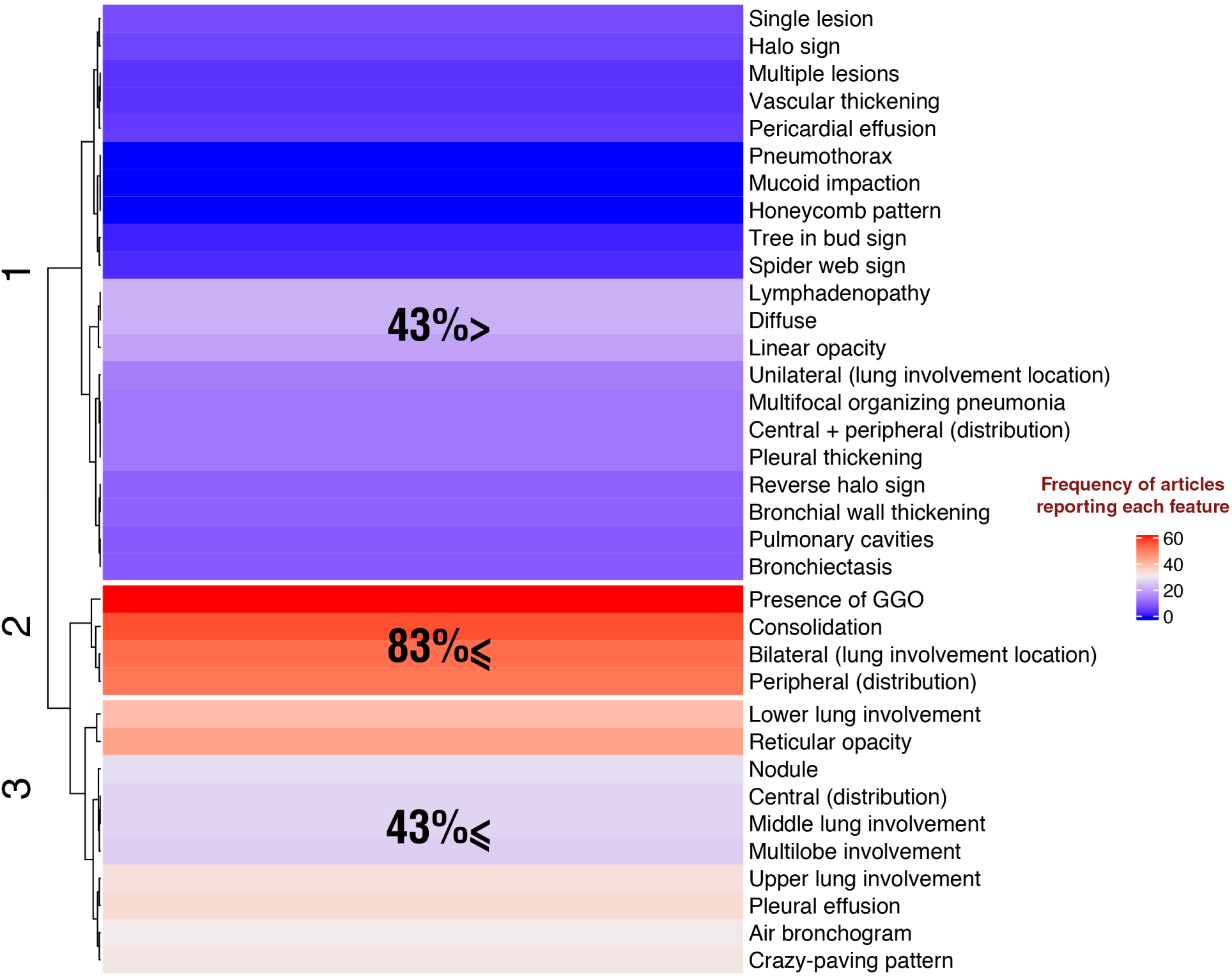
A clustergram of 35 CT imaging features of COVID-19, based on number of occurences of each feature in 60 COVID-19 related articles. Features in clusters 2 and 3 were reproted in at least 83% and 43% of studies, respectively.

### Presence of GGO, consolidation, bilateral lung involvement and peripheral distribution are the most frequent CT findings in COVID-19

The hierarchical clustering of the CT features based on their occurrences in articles was shown in figure 3. As it is evident, there are some features which are frequently reported, and therefore can be considered as typical CT features for identifying COVID-19 patients with considerable sensitivity. The most frequent features are in cluster number 3, which were reported at least in 83% of the COVID-19 studies. This cluster includes presence of GGO, consolidation, bilateral lung involvement and peripheral distribution with 100%, 92%, 85% and 83% frequency, respectively.

The pathological change of “GGO” refers to invasion of coronavirus into the bronchioles and alveolar epithelium, and their replication in the epithelial cells, causing the alveolar cavity to leak, and the alveolar wall to become inflamed, with a distribution mainly around the lung and under the pleura. GGO is a little higher density blurred image in the lungs, where the pulmonary blood vessels are visible. Also, areas of “consolidation” can be due to this fact that the pathogen invades epithelial cells, causing inflammatory thickening and swelling of the bronchial wall, but without obstructing the bronchioles. When the body reacts strongly to an inflammatory storm, large exudation occurs in the alveoli of both lungs and an area of white lung is seen on a chest CT image(11).

Both GGO and consolidation were also reported in all studies for detecting patients infected by two other strains of coronavirus family (MERS and SARS). Figure 4 shows a heat map that depicts the present/absent status of all examined CT features in all 67 studies related to COVID-19, SARS and MERS altogether. Common features between COVID-19, SARS and MERS is because of the similar pattern of lung lesion caused by different strains of coronavirus family(12).

**Figure 4.**
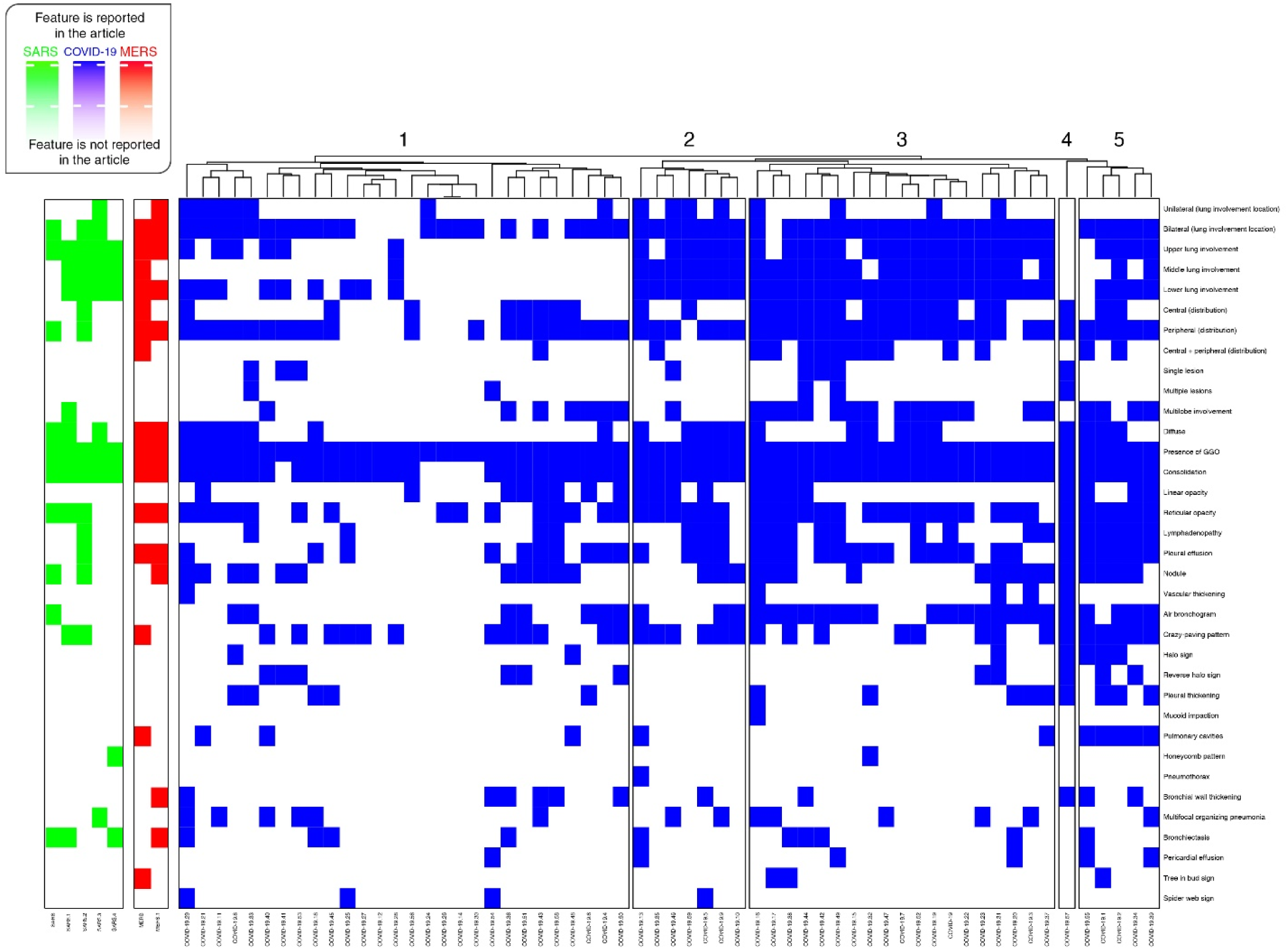
A heatmap chart to plot CT imaging features reported in each 67 articles related to COVID-19 (blue), SARS (green) and MERS (red)

The large number of COVID-19 articles reporting “bilateral lung involvement” is consistent with this observation that COVID-19 is more likely to involve both lungs on initial imaging(13). “Peripheral distribution” is another most frequently observed CT finding in these articles.

### COVID-19 specific imaging features

As Figure 4 demonstrates, several radiological presentations of COVID-19 is not much different from pneumonia associated with the other two coronaviruses. However, it seems that there are some COVID-19 specific features. Being familiar to these CT imaging features, improves our diagnostic experience and hence enhance our ability to early diagnose and combat the outbreak of COVID-19 or similar viral diseases(15). In order to find COVID-19 specific features, a discrimination score was defined (Equation 1).

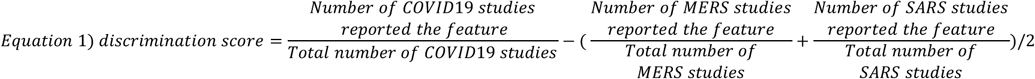

As this simple formula shows, discrimination score can range from −1 to 1. The larger the score, the more likely the feature to be COVID-19 specific. Likewise, the smaller the score, the less likely the feature to be COVID-19 specific (the more likely the feature to be SARS and MERS specific feature). Therefore, if discrimination score be near to −1 for a feature, then the feature also can be considered as a discriminative feature. Figure 5 shows the discrimination score values for all assessed features. According to figure 5, we can divide the CT features into two general categories: 1-features which can be considered as potential characteristic CT findings in patients with viral pneumonia. 2-Features which may help to discriminate COVID-19 patients from other viral pneumonia subjects.

**Figure 5.**
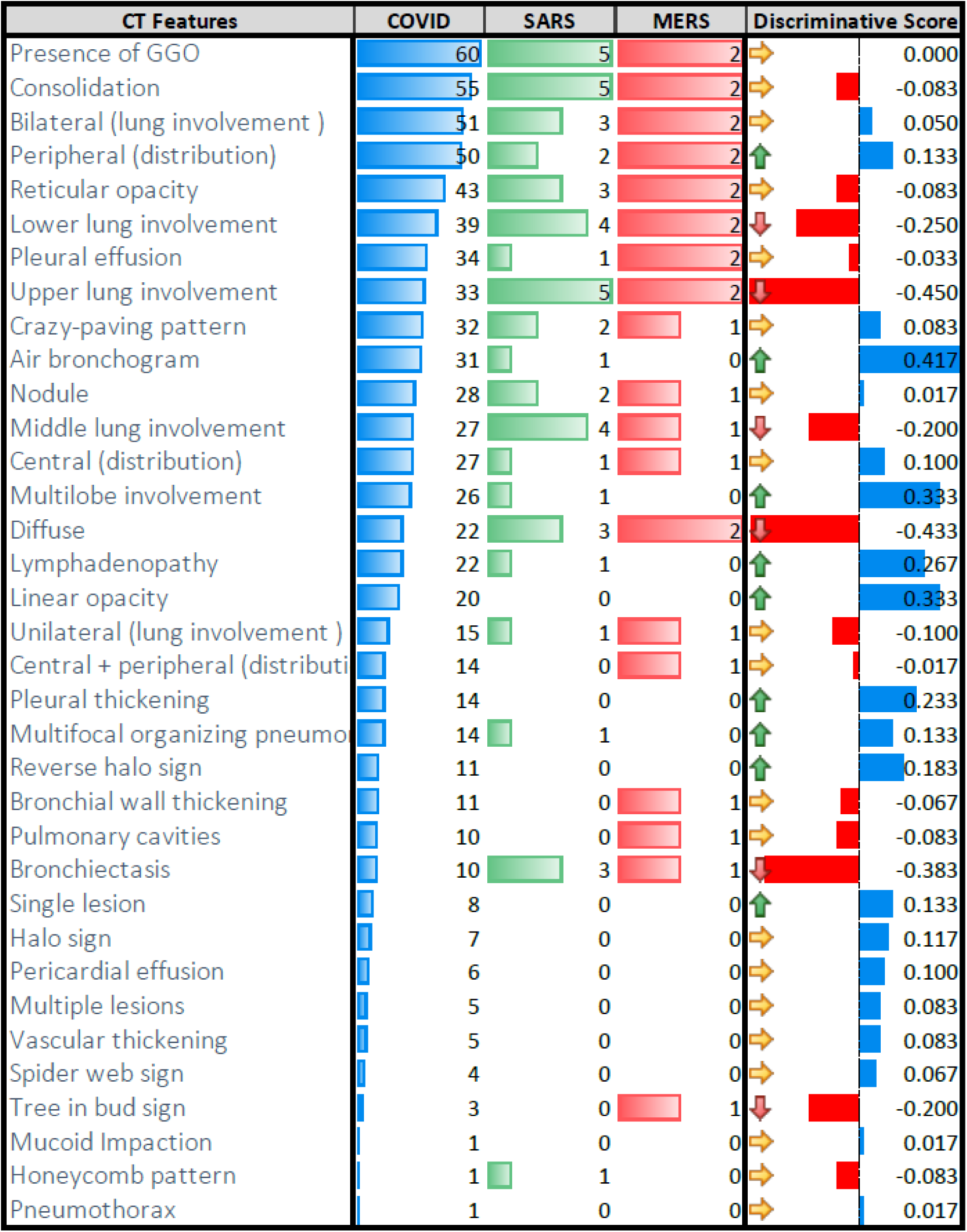
Number of articles which reported each CT imaging feature in three viral pnumonia (COVID-19, SARS, MERS). Also, last column shows the discrimination score (Equation 1) is also assigned to each feature. Larger numbers indicate COVID-19 specific features. Likewise, smaller numbers show SARS and MERS specific features.

The three CT imaging features with the highest discrimination scores are “air bronchogram”, “multi-lobe involvement” and “linear opacity”. Air bronchogram refers to the pattern of air-filled bronchi on a background of airless lung(16). When the alveoli are filled with inflammatory exudation and pulmonary consolidation, and the bronchi is inflated by positive pressure, it leads to bronchogram(17). Multi-lobe involvement is defined as when chest-radiograph shows involving more than two lobes(18). And finally, linear opacity is any opacity of approximately uniform thickness of up to 2 mm(19).

As shown in figure 5, “bilateral lung involvement” and “peripheral distribution” are the most frequently reported features among those features with a positive discrimination score. Therefore, one can conclude that these CT findings are also potential COVID-19 specific features. This is consistent with the scientific finding that COVID-19 unlike other types of pneumonia, shows a high prevalence of bilateral GGOs with a predominantly peripheral distribution on chest computed tomography images(20). In addition, according to figure 5, linear opacity, pleural thickening, reverse halo sign, single lesion, halo sign, pericardial effusion, multilobe lesions, vascular thickening, spider web sign, mucoid impaction, and pneumothorax are the 11 features with a positive discrimination score which have been reported at least once in COVID-19 articles, but have not been reported in any of the SARS and MERS articles.

Moreover, lower lung involvement, upper lung involvement, middle lung involvement, diffuse, bronchiectasis, and tree in bud sign are the 6 features with the most negative discrimination scores, that indicate their potential for being SARS and MERS specific features.

Figure 6 shows a decision tree for discriminating all 67 studies based on 35 CT features with 100% accuracy. As one can see the presence of linear opacity (the root of the tree) or pleural thickening alone can indicate that a person is infected with the COVID-19 virus. These two features are among those 11 features discussed earlier with a positive discrimination score which have not been reported in any of the studies related to SARS and MERS. On the other hand, the absence of these features, along with the absence of central and peripheral distribution, and the presence of the honeycomb pattern can indicate that a person is infected by SARS.

**Figure 6.**
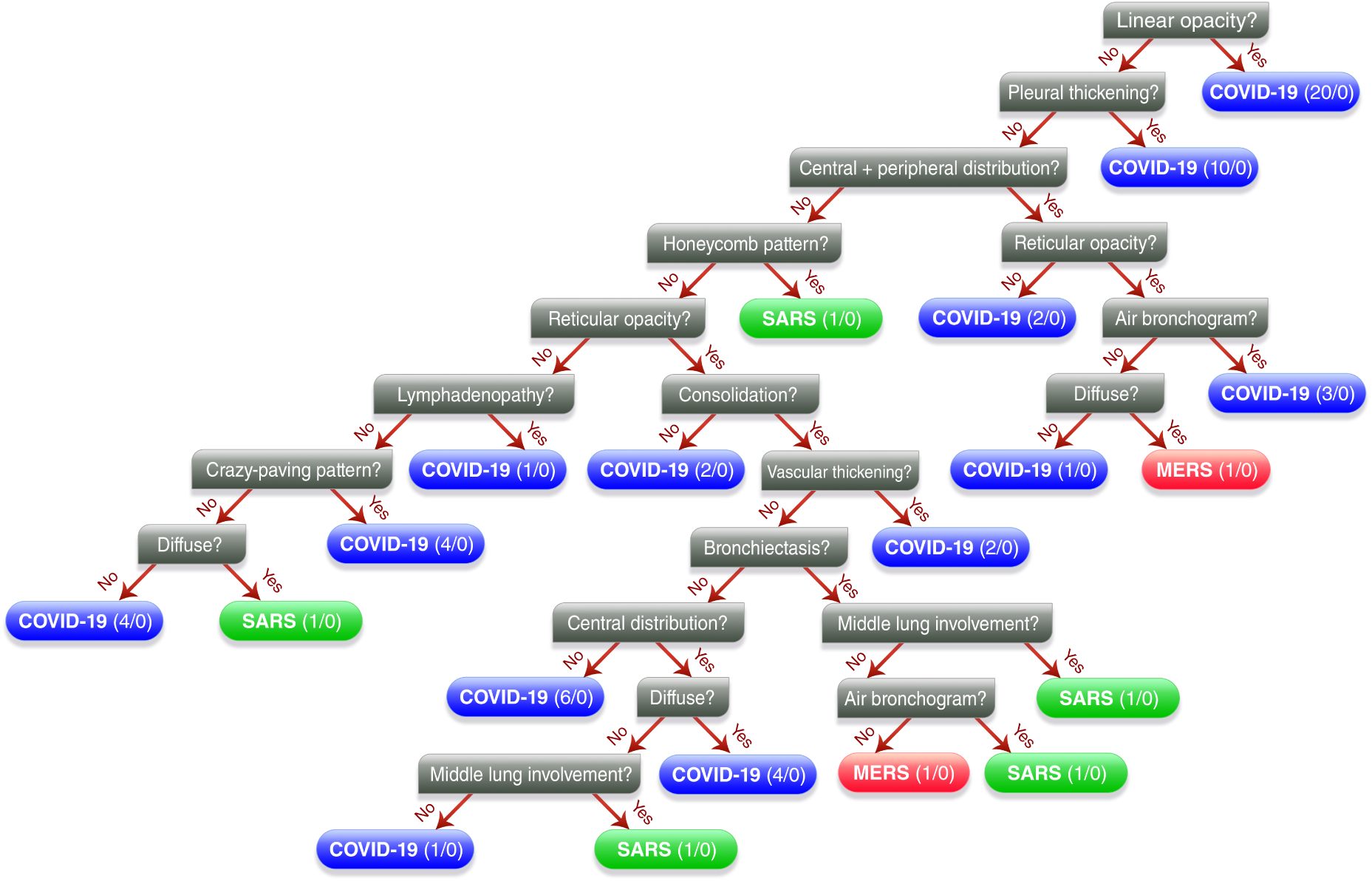
A decision tree containing a set of if-else rules to classify suspected patients into COVID-19, SARS or MERS, based on presence or absence of CT imaging features.

### Co-existed CT imaging features

By further analyses we also found clusters of co-existed features which their existence was variable among groups of studies. To be able to draw a more robust conclusion in this regard, we selected 19, out of 60 COVID-19 studies which their sample size was more than 100 (Figure 7). We did this, because very small samples undermine the internal and external validity of a study and studies with larger sample size give more reliable results and have greater precision(21).

**Figure 7.**
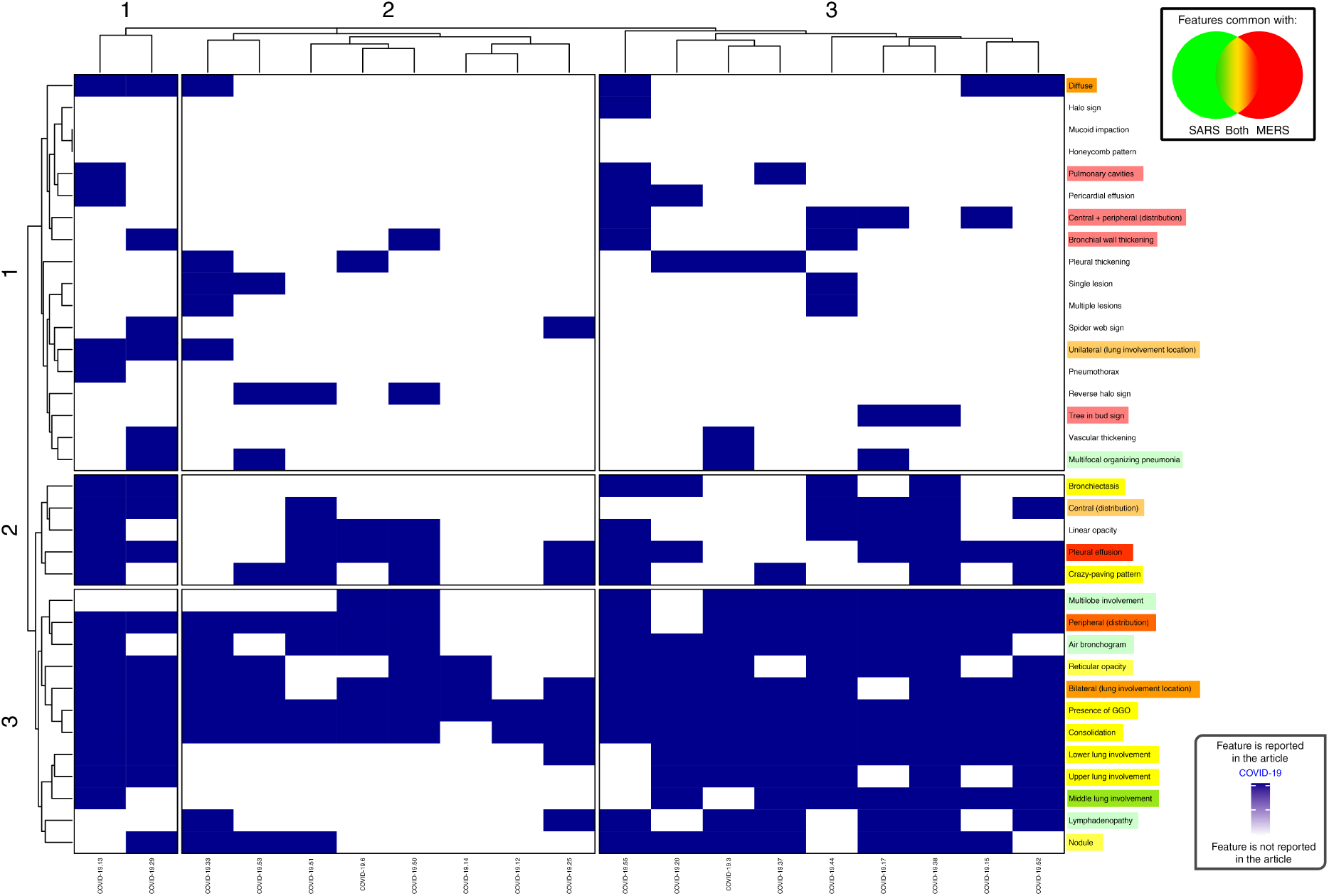
Clustergram of reported CT features in COVID-19 studies with sample size greater than 100 (19 studies). We can see 3 distinct clusters of studies based on the pattern of co-occurrences of the CT features’ absent/present. We highlighted common features between diseases in green and red, in shuch a way that the more green highlight shows the more confidence that a feature is common between COVID-19 and SARS, while the more red highlight shows the more confidence that a feature is common between COVID-19 and MERS. Also, the common features between all three diseases are highlighted as yellow, which is produced when mixing green and red in the RGB color model.

As shown in figure 7, the clustergram demonstrates that three distinct groups of studies are separated from each other based on co-occurrences of different CT findings. As we can see, pattern of co-occurrences in cluster 3 of features are well visually comparable between different clusters of studies. This cluster consists of “presence of GGO”, “consolidation”, “lower”, “upper” and “middle” lung involvement, “multi-lobe involvement”, “peripheral distribution”, “air bronchogram”, “reticular opacity”, “bilateral lung involvement”, “lymphadenopathy” and “nodule”. Most of these CT findings are common between pneumonia caused by novel corona virus and pneumonia caused by other two strains of corona virus family which their fatality rate is more(22) (23).

Another interesting result of analyzing the number of reported CT features in the COVID-19 studies is that there is a significant negative correlation (Pearson correlation coefficient: −0.3; p-value <0.02) between the number of reported features and sample size (Figure 8). It may be due to the diversity in CT findings, and not existing a large number of common features in the patients infected with COVID-19 pneumonia.

**Figure 8.**
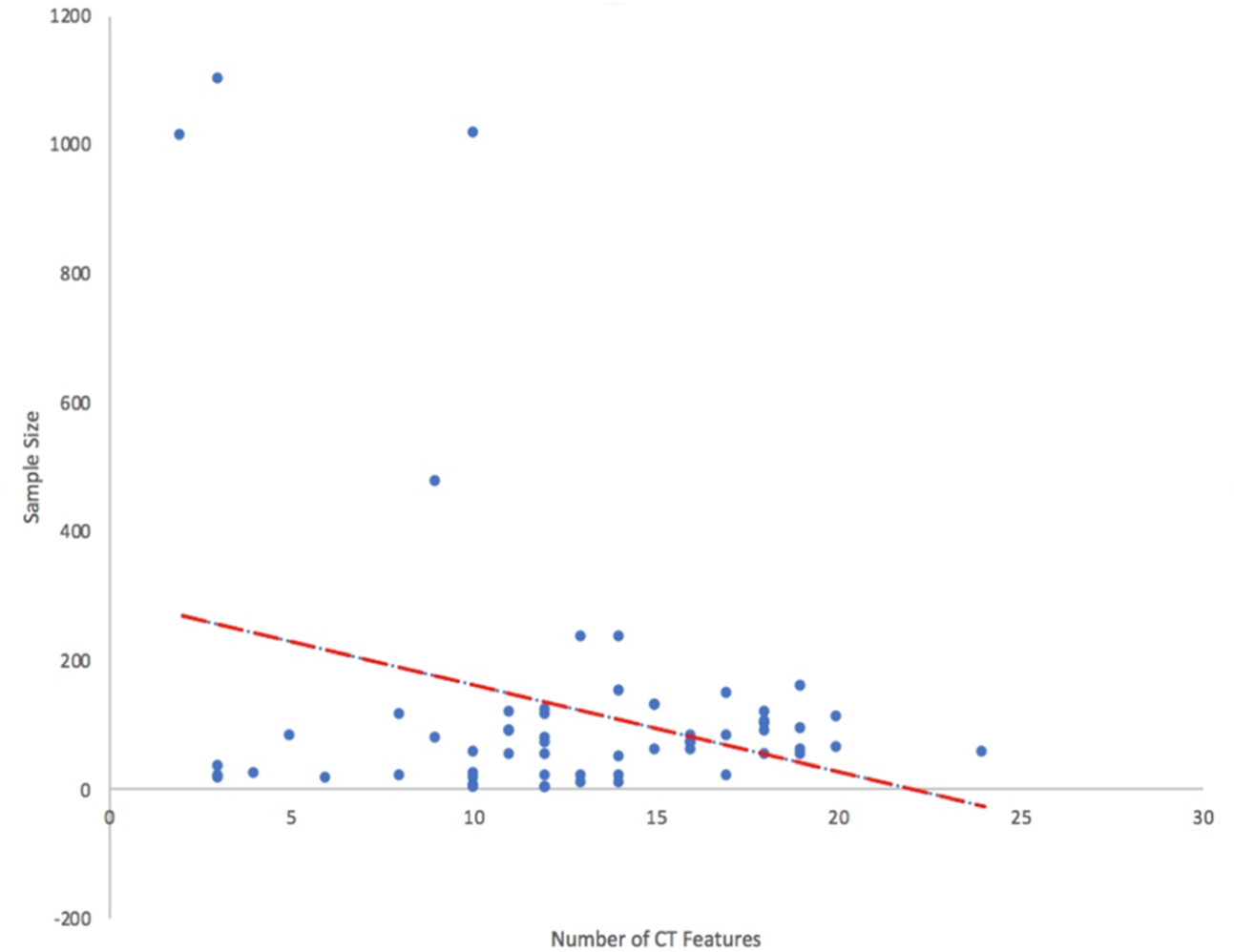
Scatter plot of sample size of COVID-19 related studies against the number of reported CT imaging features. Dotted red line is the regression line. Pearson correlation coefficient is −0.3 (p-value < 0.02).

## Conclusion

To date, a variety of CT imaging findings have been reported aimed in paving the way for diagnosis and follow-up of patients with COVID-19. It was necessary to demonstrate the most promising CT features through a reliable systematic approach for both differentiating coronaviral from non-coronaviral pneumonia and COVID-19 from the closest viral pneumonia, SARS and MERS.

Herein, we have reviewed 67 coronavirus family related studies and found the four most reported CT imaging features for detecting COVID-19: Presence of GGO, consolidation, bilateral lung involvement and peripheral distribution. Also, we identified 3 potential COVID-19 specific features: air bronchogram, multi-lobe involvement and linear opacity.

Based on the defined discrimination score, there were11 features with a positive discrimination score, but have not been reported in any of SARS and MERS studies: linear opacity, pleural thickening, reverse halo sign, single lesion, halo sign, pericardial effusion, multilobe lesions, vascular thickening, spider web sign, mucoid impaction, and pneumothorax. Also, six features had a significant negative discrimination score which may indicate that these are usually seen on radiographs of SARS and MERS patients, but not COVID-19 patients: lower lung involvement, upper lung involvement, middle lung involvement, diffuse, bronchiectasis, and tree in bud sign.

Additionally, we identified a cluster of features with co-existence within studies: presence of GGO, consolidation, lower, upper and middle lung involvement, multi-lobe involvement, peripheral distribution, air bronchogram, reticular opacity, bilateral lung involvement, lymphadenopathy and nodule.

## Data Availability

No datasets were generated during the current study

